# Deep immunophenotyping reveals associations with human blood pressure

**DOI:** 10.64898/2026.01.14.26344062

**Authors:** Evany Dinakis, Dakota Rhys-Jones, Jane Muir, Joanne A. O’Donnell, Francine Z. Marques

**Affiliations:** Hypertension Research Laboratory, Department of Pharmacology, Biomedical Discovery Institute, Faculty of Medicine, Nursing and Health Sciences, Monash University, Clayton VIC 3800, Australia; Victorian Heart Hospital, Monash University, Clayton VIC, 3168, Australia; Department of Gastroenterology, School of Translational Medicine, Monash University, Melbourne, 3000, Victoria, Australia; Baker Heart and Diabetes Institute, Melbourne, VIC 3000, Australia

## Abstract

Preclinical models have been instrumental in defining the immunological mechanisms underlying hypertension, yet human studies remain comparatively limited, leaving a major gap in translational understanding. To address this, we investigated immune cell dynamics in human hypertension by profiling peripheral blood mononuclear cells from 48 normotensive and hypertensive participants, characterised using ambulatory blood pressure (BP) monitoring. High□dimensional spectral flow cytometry revealed strong positive correlations between night□time systolic BP and multiple CD8□T□cell subsets. Hypertensive participants also exhibited significantly elevated CD8□T cell subsets, a pattern that persisted even among those receiving anti□hypertensive medication with subsequently controlled BP. These findings suggest that BP control does not necessarily equate to biological control of hypertension and highlight a distinct CD8□T cell–driven signature that differentiates individuals with normal BP and hypertension.

---

A growing body of evidence indicates that both the innate and adaptive immune systems play a significant role in the pathogenesis and progression of hypertension, arguably making it an unconventional inflammatory condition (1). Experimental models have consistently demonstrated the central role of adaptive immunity; for example, mice lacking both B and T cells exhibit markedly blunted blood pressure (BP) elevations and reduced vascular injury, underscoring the contribution of lymphocytes in BP regulation (2). Notably, reconstitution of T cells restored hypertensive responses, highlighting their causal role (2). In parallel, T cell– derived cytokines such as interleukin-17 (IL□17) drive vascular inflammation and end□organ damage (3), further establishing T cells as key mediators of hypertensive pathology. Innate immune pathways similarly shape the inflammatory milieu that supports T cell priming and sustained activation (1), creating a feed□forward cycle that is primarily centred on T cell–mediated responses. However, most of this mechanistic understanding arises from preclinical models of hypertension. Whether these immune mechanisms operate similarly in humans remains less clear. Here, we aimed to comprehensively characterise the peripheral immune system in human hypertension. Using peripheral blood mononuclear cells isolated from 48 participants characterised with ambulatory BP monitoring, we quantified 121 immune cell populations and subtypes, defined by their activation status and cytokine expression profiles, using spectral flow cytometry (4) (Figure A).

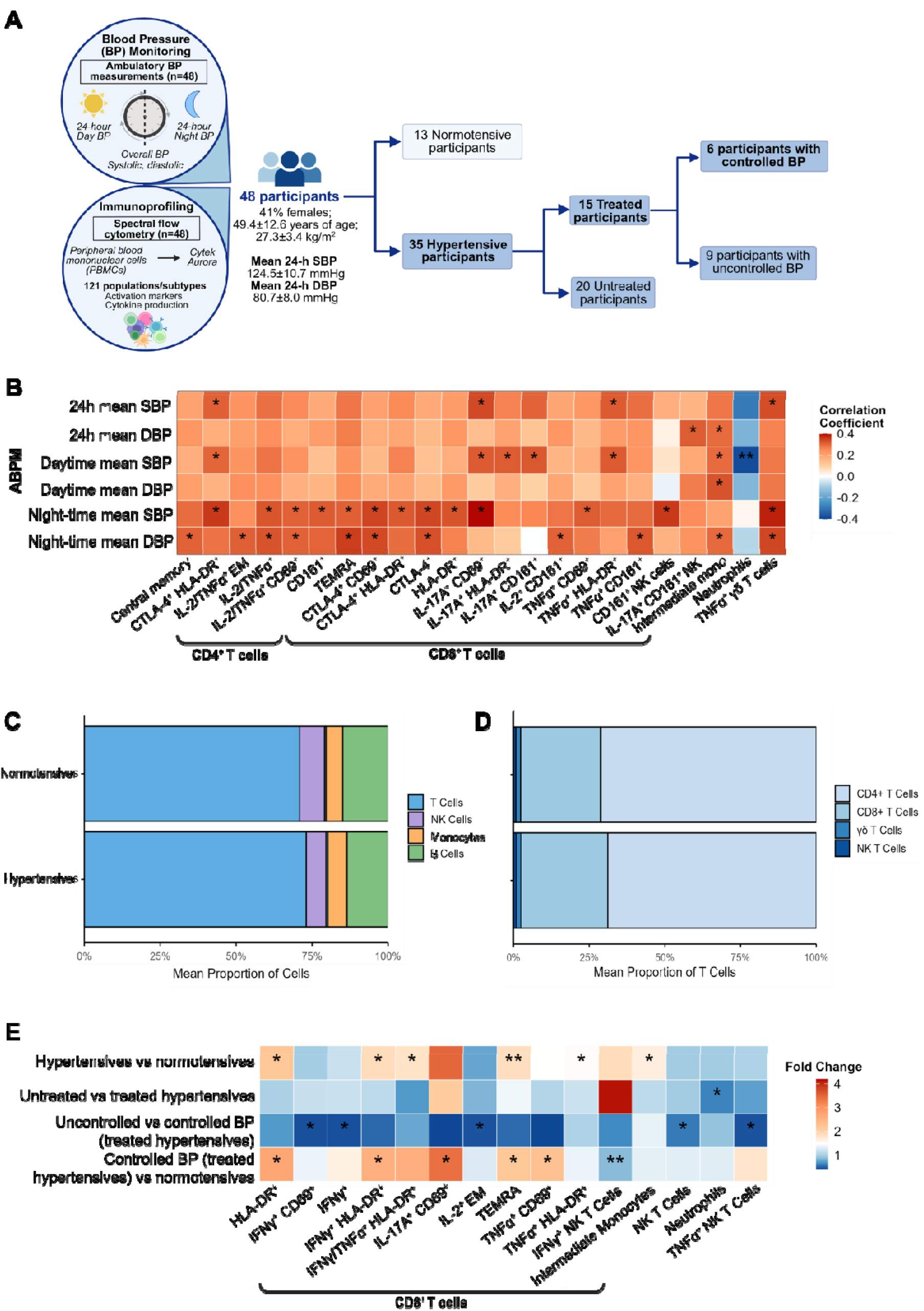
**A**, The pH of Intestines and Blood-pressure Regulation (pHibre) study was approved by the Monash University Human Research Ethics Committee (Study ID: 23336) and conducted in accordance with the Declaration of Helsinki, detailed elsewhere (4). All participants provided written consent. The study was registered under the Australian New Zealand Clinical Trials Registry (ACTRN12620000284965). Spectral flow cytometry was performed on peripheral blood mononuclear cells (PBMCs) isolated from fasted whole blood (n=48). Cohort characteristics are reported as mean□±□SD. **B**, Spearman’s rank correlations as a heatmap between ambulatory blood pressure measurements (ABPM) including 24-hour, day- and night-time systolic BP (SBP) and diastolic BP (DBP), and the peripheral immune system. Mean proportion of **C**, major immune cell populations and **D**, major T cell subsets in normotensive and hypertensive participants. **E**, Heatmap depicting fold□change values derived from Mann–Whitney tests comparing hypertensive and normotensive participants, untreated and treated hypertensives, treated hypertensives with uncontrolled and controlled BP, and controlled hypertensives and normotensive participants. Normality was assessed using the Shapiro-Wilk test. Outliers were removed using the 1.5× interquartile range method. Corrections for multiple comparisons were performed using the Benjamini– Hochberg false discovery rate method (all q>0.05, data not shown). The colour gradient represents the strength and direction of association (red = positive, blue = negative). P-values depicted in heatmaps: ^*^P<0.05, ^**^P<0.01. EM, effector memory; TEMRA, terminally differentiated effector memory T cells; NK, natural killer; mono, monocytes.

Spearman’s rank correlation analyses revealed 38 significant associations between the peripheral immune system and 24-hour, day- and night-time systolic and diastolic BP (SBP and DBP, respectively). These associations were predominantly positive, driven largely by early (CD69^+^) and late-activated (HLA-DR^+^) CD8^+^ T cells, as well as pro-inflammatory cytokine-producing subsets, including IL-17A- and TNFα-producing CD8^+^ T cells (Figure B). Thirteen of these were linked to night-time SBP, with IL-17A-producing CD69^+^ CD8^+^ T cells (*P*=0.011) and TNFα-producing γδ T cells (*P*=0.016) showing the strongest relationships (Figure B). Night□time SBP is increasingly recognised as a more stable and physiologically representative measure of true BP, as it is less influenced by behavioural and environmental fluctuations (5). Consequently, the associations observed at night□time are especially compelling, likely reflecting more robust and biologically meaningful immune– cardiovascular interactions.

Participants were classified as normotensive (n=13) or hypertensive (n=35) using ambulatory hypertension thresholds (day and/or night SBP/DBP ≥135/85 or 120/75 mmHg, respectively). Overall, the proportions of major immune cell populations and T cell subsets were comparable, with T cells constituting the dominant population in both groups (normotensives: 70.9% vs hypertensives: 73.1%, Figures C and D). However, the majority of differences were driven primarily by CD8□ T cell subsets (Figure E). For example, hypertensive participants had a 2.2□fold increase in HLA□DR□ CD8□ T cells and a 1.9-fold increase in IFNγ□producing HLA□DR□ CD8□ T cells. The most statistically significant difference was observed in terminal effector memory (TEMRA) CD8□ T cells (mean±SEM, normal BP: 7,633.42±2,146.93 vs high BP: 14,032.64±1,719.88, *P*=0.032, Figure E).

Participants with hypertension were stratified into untreated (n=20) and treated (i.e., receiving anti-hypertensive medication, n=15) groups. Treated hypertensive participants exhibited higher neutrophil counts (treated: 977.60±130.94 vs untreated: 591.06±88.43, *P*=0.016, Figure E). However, not all treated patients achieve adequate BP control. To capture this distinction, treated hypertensive participants were further stratified into uncontrolled (n=9) and controlled (n=6) BP groups. Relative to treated hypertensive participants with controlled BP, the uncontrolled group exhibited a −2.5□fold decrease in both IFNγ□producing and proliferating (IL-2^+^) CD8□ T cells (Figure E). Moreover, those with uncontrolled BP also had lower natural killer (NK) T cells (uncontrolled BP: 2,355.67±767.74 vs controlled BP: 4,095.0±656.45, *P*=0.050) and TNFα-producing NK T cells (uncontrolled BP: 263.50±66.48 vs controlled BP: 609.33±125.93, *P*=0.013, Figure E). Finally, a sensitivity analysis showed that participants with hypertension, irrespective of treatment or BP control, still exhibited five elevated CD8□ T cell populations relative to participants with normal BP. Notably, three of these populations overlapped with differences observed in the primary comparison between hypertensive and normotensive participants (i.e., HLA-DR^+^, IFNγ-producing HLA-DR^+^ and TEMRA CD8^+^ T cells, Figure E).

We acknowledge that this study has limitations, most notably the small cohort size, which becomes even more restricted when participants are stratified into sub□groups. Nevertheless, this is the largest study to date to provide a detailed characterisation of immune cell populations and their functional states, including activation and cytokine production, thereby yielding deeper insights into peripheral immune mechanisms associated with hypertension. Importantly, the peripheral immune phenotypes observed here likely mirror underlying tissue□level inflammation and T cell–driven vascular injury, consistent with mechanisms described in preclinical models(2, 3), suggesting that circulating immune signatures may serve as accessible indicators of deeper immunopathology.

In conclusion, our findings indicate that BP and hypertension are associated with changes to the peripheral immune system in humans. However, neither BP□lowering medication nor achieving BP control fully restores immune balance in hypertension. This suggests that hypertension itself is the underlying driver of the CD8□ T cell–dominated inflammatory profile.

## Data Availability

All data produced in the present study are available upon reasonable request to the authors

## Acknowledgements

This study was supported by a National Heart Foundation Vanguard Grant (102927) and a National Health & Medical Research Council of Australia (NHMRC) Ideas Grant (GTN2017382). F.Z.M. is supported by a Senior Medical Research Fellowship from the Sylvia and Charles Viertel Charitable Foundation, a National Heart Foundation Future Leader Fellowship (105663), and a National Health & Medical Research Council (NHMRC) Emerging Leader Fellowship (GNT2017382). J.A.O. is supported by a NHMRC Fellowship (GNT1124288). The authors thank Monash University FlowCore, the Monash Statistical Consulting Service, the Monash eResearch capabilities for providing access to M3 servers.

